# Clinical and Economic impact of updated Fall 2023 COVID-19 vaccines in the Immunocompromised Population in Canada

**DOI:** 10.1101/2023.11.10.23298369

**Authors:** Amy Lee, Kavisha Jayasundara, Michele Kohli, Michael Maschio, Kelly Fust, Keya Joshi, Nicolas van de Velde, Ekkehard Beck

## Abstract

Background

Immunocompromised (IC) individuals are at increased risk of COVID-19 infection-related severe outcomes. Moderna and Pfizer-BioNTech COVID-19 mRNA vaccines are available in Canada, and differences in vaccine effectiveness (VE) have been found between the two in IC individuals. The objective of this analysis was to compare the clinical and economic impact of a Moderna XBB.1.5 updated COVID-19 mRNA Fall 2023 vaccine to a Pfizer-BioNTech XBB.1.5 updated COVID-19 mRNA Fall 2023 vaccine in Canadian IC individuals aged ≥18 years.

**Methods:** A static decision-analytic model estimated the number of COVID-19 infections, hospitalizations, deaths, and resulting quality-adjusted life years (QALYs) over a one-year time horizon (September 2023-August 2024) in the Canadian IC adult population (n=894,580). Costs associated with COVID-19 infection were estimated from health care and societal perspectives. The predicted VE of the updated Moderna vaccine was based on prior variant versions, which were well-matched to the circulating variant. Pfizer-BioNTech VE was calculated based on a meta-analysis of comparative effectiveness between both vaccines (relative risk for Moderna vaccine: infection=0.85 [95%CI 0.75-0.97], hospitalization=0.88 [95%CI 0.79-0.97]). The model combined VE estimates with COVID-19 incidence and probability of COVID-19 related severe outcomes. Sensitivity analyses tested the impact of uncertainty surrounding incidence, hospitalization and mortality rates, costs, and QALYs.

**Results:** Given the expected higher VE against infection and hospitalizations with the Moderna Fall 2023 vaccine, its use is predicted to prevent an additional 2,411 infections (3.6%), 275 hospitalizations (3.7%), and 47 deaths (4.0%) compared to the Pfizer-BioNTech Fall 2023 vaccine, resulting in 330 QALYs gained, and savings of $7.4M in infection treatment costs, and $0.9M in productivity loss costs. Results were most sensitive to variations in VE parameters, specifically the relative risk of infection and hospitalizations between the vaccines, and waning rates.

**Conclusions:** If the Moderna and Pfizer-BioNTech Fall 2023 vaccines protect against infection and hospitalizations similar to previous vaccines, using the Moderna Fall 2023 vaccine would result in substantial public health benefits in IC individuals, as well as provide health care and societal cost savings.

## INTRODUCTION

On May 5, 2023, the World Health Organization (WHO) declared that COVID-19 is no longer considered a public health emergency, but an ongoing health issue.^1^ It recommended that vaccination efforts continue, particularly in high-priority groups. As the SARS-CoV-2 virus continues to evolve, there is an emphasis on updating the vaccines to target the current variants of concern. On September 12, 2023, the Canadian National Advisory Committee on Immunization (NACI) amended their guidance on the use of COVID-19 vaccines in the Fall of 2023.^2^

The updated Canadian guidance advises the use of an XBB.1.5-containing COVID-19 vaccine, with particular importance placed on those at increased risk of COVID-19 severe disease, such as those with underlying medical conditions, including those with primary immunodeficiency diseases, malignancies or who use immunosuppressive medications.^3^

Compromised immune systems can be due to genetic defects or conditions that suppress the immune system,^4^ such as solid tumours or hematologic malignancies, solid organ or hematopoietic stem cell transplants, chronic kidney disease on dialysis, human immunodeficiency virus (HIV), and chronic use of immunosuppressive or immunomodulatory medications.^5^ These individuals are considered to be moderately to severely immunocompromised by the Canadian National Advisory Committee on Immunization (NACI),^5^ and are at greater risk of hospitalizations, and mortality from infectious diseases, including COVID-19.^4,6–9^ It is therefore important to increase their immune response against SARS-CoV-2 through vaccination.

The protective response mounted by immunocompromised (IC) individuals to the COVID-19 vaccinations is decreased compared to their immunocompetent counterparts. In a prospective cohort study conducted at Kaiser Permanente Southern California, Tseng et al., (2023) found the adjusted vaccine effectiveness (VE) for the general population of the Moderna BA.4/BA.5 bivalent vaccine (mRNA-1273.222) against hospitalization was 82.8%, but reduced to 71.8% in those that were IC.^10^ Additionally, similar to the general population,^11–15^ VE has been found to differ between the two mRNA COVID-19 vaccines in the IC population. Although both Moderna and Pfizer-BioNTech COVID-19 vaccines utilize mRNA technology, the formulation, including the type of nanoparticles used for the mRNA delivery system, and dosage of each vaccine differ.^16–19^ A systematic literature review and meta-analysis based on 17 studies by Wang et al., (2023) found that in the IC population, the Moderna mRNA-1273 vaccine to be associated with significantly lower risk of COVID-19 infection, hospitalizations, and deaths compared to the Pfizer-BioNTech BNT162b2 vaccine (relative risk for infection = 0.85 [95% CI 0.75-0.97]; hospitalization = 0.88 [95% CI 0.79-0.97]).^20^

Given the approval of the recently updated COVID-19 vaccines in Canada for both Moderna and Pfizer-BioNTech,^21,22^ the question arises on whether and to which extend there may be clinical or economic differences between the use of these two mRNA vaccines in this vulnerable IC population. The objective of this analysis was to estimate the clinical and economic impact of a Moderna XBB.1.5 updated COVID-19 mRNA Fall 2023 vaccine (Moderna Fall 2023 vaccine) compared to a Pfizer-BioNTech XBB.1.5 updated COVID-19 mRNA Fall 2023 vaccine (Pfizer-BioNTech Fall 2023 vaccine) in IC individuals aged 18 years over a one-year time horizon from September 2023 to August 2024.

## METHODS

### Overview

A static decision analytic model was developed in Microsoft Excel to estimate the number of infections, hospitalizations, deaths, and costs associated with COVID-19 treatment over the course of one year (September 2023-August 2024) for the Canadian population. The model combined estimates of the incidence of COVID-19 in Canada, the effectiveness of updated COVID-19 vaccinations and the probability of severe outcomes from COVID-19 infections. The economic costs were estimated from both health care system and societal perspectives. Outcomes estimated included the number of COVID-19 infections, hospitalization and deaths as well as associated quality-adjusted life-years (QALYs) and costs.

The target population of this analysis was moderately to severely IC individuals in Canada aged ≥18 years considered to be at increased risk of severe disease from COVID-19 as defined by the National Advisory Committee on Immunization (NACI).^23^

A Markov model with tunnel states was used to track the monthly COVID-19 vaccination status of the target population, over a one-year analytic time horizon. At the start of the time horizon (September 2023), the 2023 Canadian IC population aged ≥18 years was assigned to the following historical vaccine coverage strata based on the highest level of prior COVID-19 vaccine received: did not receive primary series; received primary series; received first booster; received second booster. For historical vaccination coverage in the IC population, 3 doses (2 doses for Ad26.COV2.S) were considered primary series.^5^ Additional doses were considered boosters. Data on COVID-19 vaccine coverage in the IC population up to August 13, 2023 were obtained from the Institute for Clinical Evaluative Sciences (ICES), which provided number of individuals in priority groups vaccinated by dose, and risk conditions.^24^ Risk conditions that matched the Canadian Immunization Guide COVID-19 vaccine’s criteria for moderate/severe IC were included.^5^ The percentage of the IC population by age (18-59 years, ≥60 years) that received primary series, 1 booster, or 2 boosters as their highest level of vaccination was calculated by dividing the total number of IC individuals that received each level of vaccination by the total number of IC individuals. The distribution of individuals, by age, and vaccination strata, are displayed in Table 1.

**Table 1.** Historical coverage in immunocompromised individuals, by highest level of vaccination and age.

| Age | Did not receive primary series | Received primary series only | Received first booster | Received second booster |
| --- | --- | --- | --- | --- |
| 18-59 years | 41.9% | 32.9% | 17.9% | 7.3% |
| 60+ years | 14.7% | 21.2% | 23.0% | 41.1% |

Individuals who had completed at least primary series vaccination were able to move to the Fall 2023 vaccination strata. The model simulation was run twice: once where those in the Fall 2023 vaccination strata receiving the Moderna Fall 20203 vaccine and once where they receive the Pfizer-BioNTech Fall 2023 vaccine. The outcomes from each model run were compared.

The maximum Fall 2023 uptake was age-group specific and assumed to be the same as that achieved by the general Canadian population for the COVID-19 second booster, as these were the most complete data available.^25^ Roll-out of the second booster was extremely staggered, given timing of the second booster and 6 month recommended time interval between doses, therefore uptake patterns from a shorter vaccine campaign was needed. Uptake was assumed to occur over five months (September 2023-January 2024), as observed during the 2021-2022 seasonal influenza vaccination campaign.^26^ Furthermore, the proportion of the total COVID-19 vaccinations that were administered each month was based on the monthly uptake rates observed during the 2021-2022 influenza campaign.^26^ The final uptake pattern by month and age is displayed in Figure 1. Individuals receiving the Fall 2023 vaccine are drawn from each vaccination strata proportionally. Optimistic scenario analyses were included where the uptake in the 80+ years age group was increased from 73% to 80% (increase of 9%) and 95% (increase of 30%), with all other age group values adjusted up by the same multiplier.

**Figure 1.**
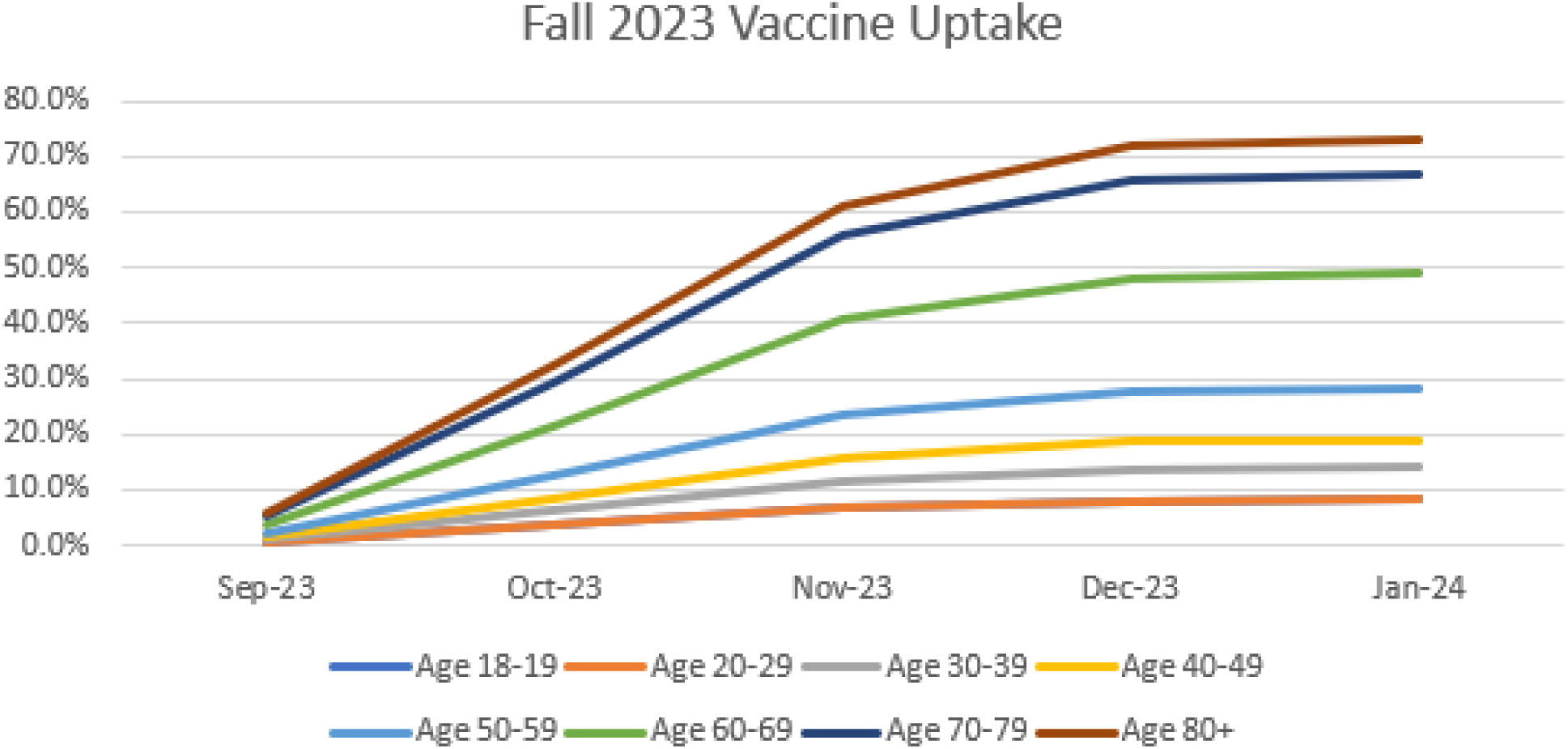
Fall 2023 COVID-19 vaccine uptake, by age group.

The cohort begins the model in the Well health state (Figure 2). Each month, individuals are at risk of COVID-19 symptomatic infection, which is dependent on infection incidence. The risk is decreased by prior vaccination status, as well as whether they received the Fall 2023 vaccine. If the individual develops a COVID-19 symptomatic infection, they move through an infection consequence decision tree (Figure 3, described in detail further below), which calculates subsequent costs and clinical outcomes of infection. Those with infections can remain in the Well health state (shown as Recovered in the consequence decision tree), or transition to the Dead health state (shown as In-Hospital Mortality in the decision tree). Death in the model is only due to COVID-19 due to the short time horizon. All-cause mortality is not included.

**Figure 2.**
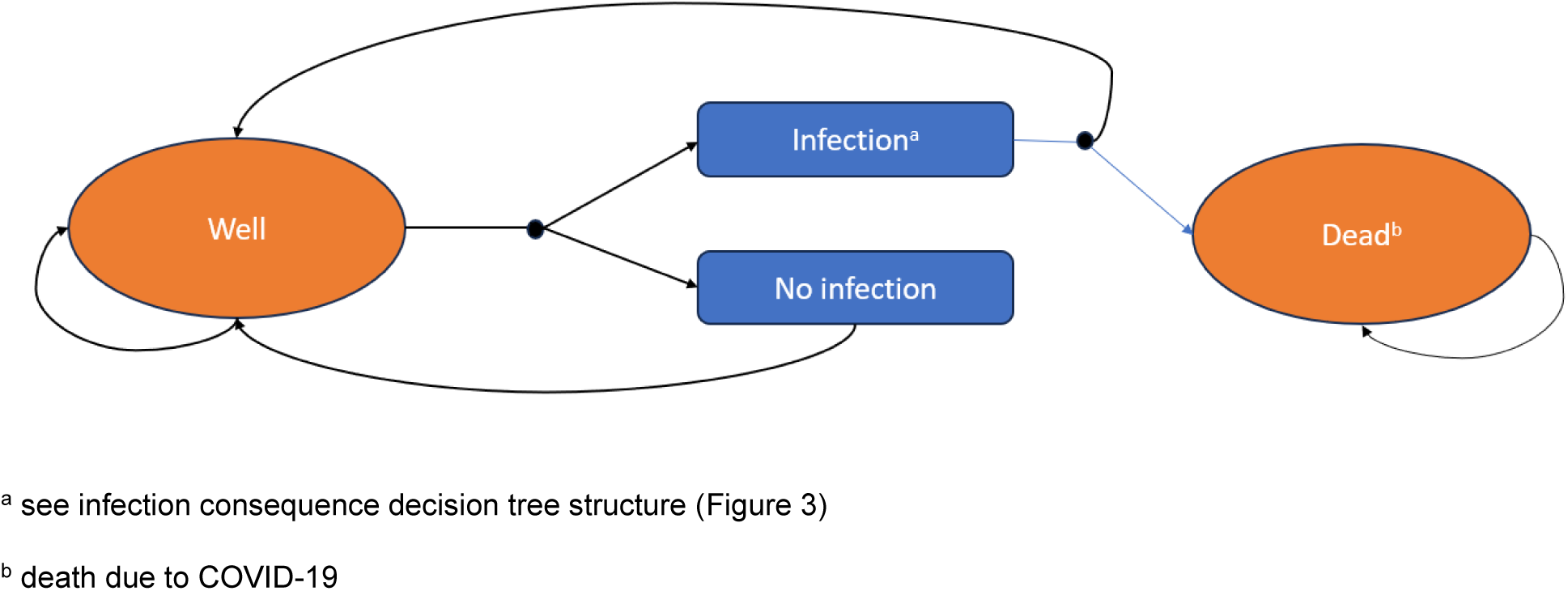
Model structure.

**Figure 3.**
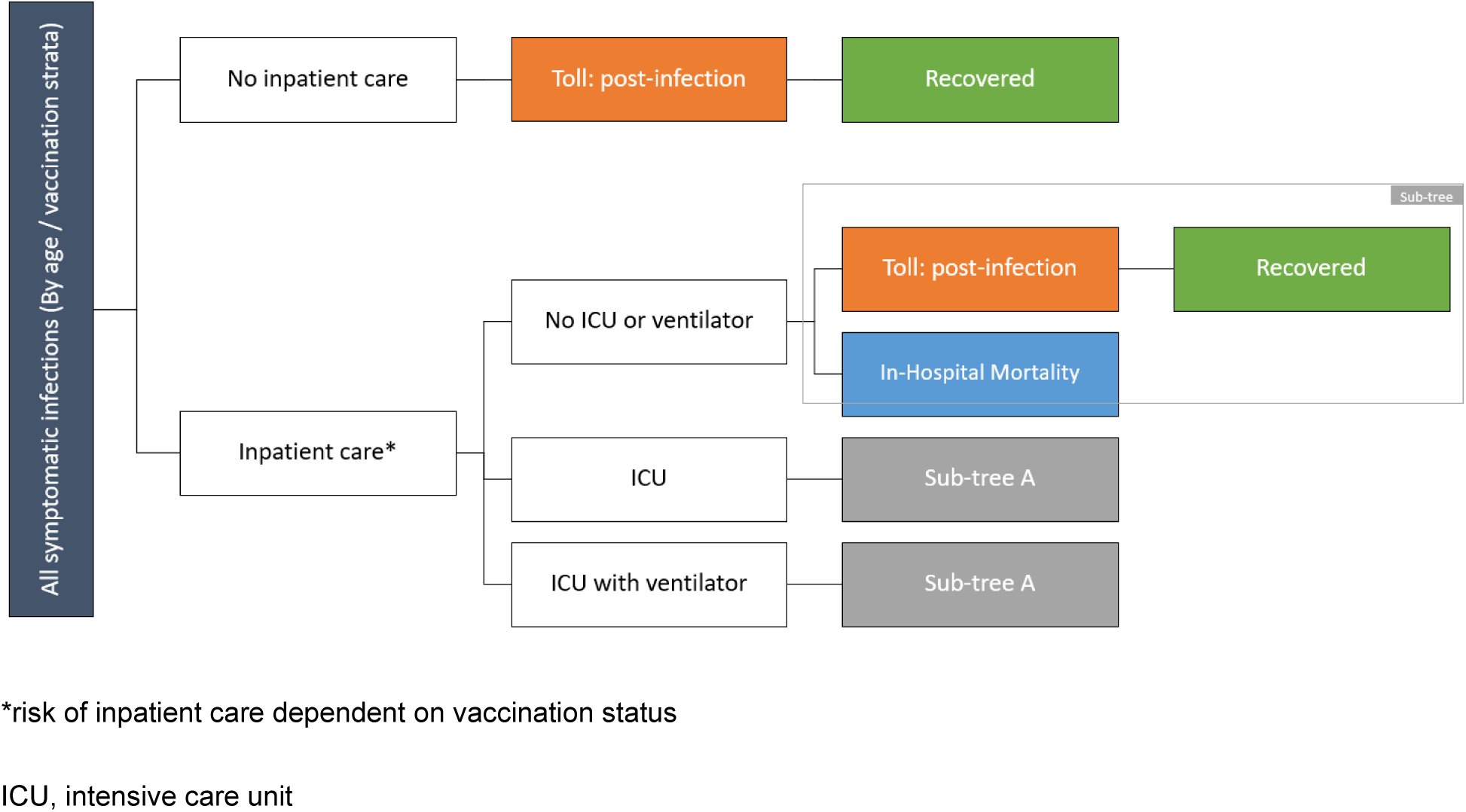
Infection consequence decision tree structure.

### Population size

An IC population size of 894,580 was included in the analysis. This was estimated, by age group, based on a US-based survey to determine the prevalence of immunocompromised individuals,^27^ as well as a Statistics Canada survey conducted near the start of the COVID-19 pandemic to determine the breakdown of the population by age.^28^ Further details are provided in the Supplementary Material.

### Incidence

As the true incidence of COVID-19 infection in Canada is underestimated due to the lack of mandatory testing and reporting, the infection incidence was calculated using Public Health Agency of Canada (PHAC) COVID-19 hospitalization data for the general population for September 2022 to August 2023.^29^ Data collection for July and August 2023 were incomplete and therefore, the ratio of the rates from June 2022 to June 2023 was applied to the rates from July and August 2022 to approximate the hospitalization rates for July and August 2023.

The symptomatic infection incidence required to result in the predicted monthly rate of hospitalizations was then back-calculated. To do this, calibration was performed using a version of the Markov model populated with general population data, with the calibration target set to the expected number of COVID-19 hospitalization data based on the data from PHAC. This method estimated the monthly symptomatic infection incidence for September 2022 to August 2023. In the base case analysis, it was then assumed that the COVID-19 incidence was the same for September 2023-August 2024 as the prior year.

The projected monthly incidence rates of COVID-19 infection for September 2023-August 2024 and additional details are provided in the Supplementary Material.

### Residual vaccine effectiveness

Residual VE is necessary to estimate the level of protection in the population entering the model at the start of the analytic time horizon (September 1, 2023) due to prior vaccinations. This is calculated using the timing of the prior boosters, VE of primary series and boosters against the dominant variant at time of administration, and the historical coverage in the IC population.

The VE of primary series and monovalent boosters against Omicron BA.1/BA.2 for infection and hospitalizations in the general population were obtained from the meta-analysis by Pratama et al. (2022).^30^ As there is a mix of primary series vaccines, the VE estimates for all vaccines were used. For the VE of booster vaccination, results for mRNA vaccines were used. Primary and booster monovalent VE against hospitalizations were further adjusted to account for the reduced VE observed in the IC population and due to the emergence of BA.4/5 by using the ratio between monovalent against omicron in the general population, and bivalent against BA.4/BA.5 in the IC population.

Bivalent boosters (assumed to be first booster for some, and second booster for all in the IC population) were available during the period of BA.4/5 dominance. The VE for bivalent boosters against hospitalization in the IC population was obtained from a Kaiser Permanente study of the Moderna bivalent vaccine (71.8%, 95% CI 48.8%-84.5%).^10^ As VE against infection was not available for the IC population, it was assumed to be equivalent to monovalent vaccine against Omicron (57.1% from Pratama et al., 2022^30,31^) for the general population.

Monthly VE waning for primary series and boosters for both infection (4.8%) and hospitalization (1.4%) were assumed to be the same monovalent booster waning rates estimated in the meta-analysis by Higdon et al., (2022).^31^

Timing of prior boosters was obtained from coverage by week and age data from Public Health Ontario.^32^ The month when 50% of vaccine were administered for primary series (Dec 2021), first booster (May 2022 for those who received monovalent booster, November 2022 for those who received bivalent booster), and second booster (November 2022, bivalent) for the oldest age group was used as a proxy for vaccine administration date for each level of vaccination (primary series, boosters).^32^ The timing of vaccine administration differed among age groups but this simplifying assumption was adopted because the older age groups had significantly higher prior coverage levels compared to younger age groups. Some IC individuals received monovalent and some received bivalent vaccines as their second booster. Due to the difference in VE between these two vaccine types, and timing of when administered, the residual VE of both were calculated, then weighed by proportion that received each (60% received monovalent, and 40% received bivalent as their second booster).^33^ Those that received the second booster mainly consisted of individuals aged 60+ that received a monovalent vaccine as their first booster earlier in the year. It was assumed that the IC population completed the same level of vaccinations similar to the general population, with the adjustment for the additional dose during primary series for the IC population. The residual VE at start of the analytic time horizon is shown in Table 2.

**Table 2.** Residual vaccine effectiveness in the immunocompromised population at start of analytic time horizon.

|  | **Infection** | **Severe** |
| --- | --- | --- |
| Completed primary series | 0.0% | 21.5% |
| Completed first booster | 3.7% | 53.2% |
| Completed second booster | 9.3% | 58.1% |

### Vaccine effectiveness of the Moderna and Pfizer-BioNTech Fall 2023 vaccines

Real world effectiveness on the Moderna and Pfizer-BioNTech Fall 2023 vaccines are not yet available. Therefore, in the base case analysis, the vaccines are assumed to be well-matched to the current variant, and are assumed to protect at the same level against the circulating dominant variant as the bivalent booster (mRNA-1273.222 and BNT126b2 [WT/ OMI BA.4/BA.5]) protected against BA.4/BA.5. Therefore, like the bivalent booster values used to calculate residual effectiveness, the initial VE of the Moderna Fall 2023 vaccine was set to 71.8% for hospitalization based on the Kaiser Permanente study of the Moderna bivalent vaccine,^15^ and 57.1% for infection, based on the estimate for monovalent vaccine against Omicron from Pratama et al.^30^ To approximate the VE of a well-matched Pfizer-BioNTech Fall 2023 vaccine, the relative risk of hospitalization and infection in the IC population for the Moderna bivalent booster compared to the Pfizer-BioNTech bivalent booster (relative risk [RR] = 0.88 and 0.85, respectively) from Wang et al., (2023) was applied to the assumed VEs for the Moderna Fall 2023 vaccine.^20^ This resulted in estimated initial VEs of 68.0% and 49.6%. The waning of the Fall 2023 vaccines were assumed to be the same as the monovalent booster waning rates estimated in the meta-analysis by Higdon et al., (2022).^31^ VE inputs for the base case and sensitivity analyses are summarized in Table 3.

**Table 3.** Base case and sensitivity analyses vaccine effectiveness inputs.

| Description of analysis | Moderna Fall 2023 vaccine |  |  |  | Pfizer-BioNTech Fall 2023 vaccine |  |  |  |
| --- | --- | --- | --- | --- | --- | --- | --- | --- |
|  | Infection |  | Hospitalization |  | Infection |  | Hospitalization |  |
|  | Initial<br>VE | Waning | Initial<br>VE | Waning | Initial<br>VE | Waning | Initial<br>VE | Waning |
| Base case analysis | 57.1% | 4.8% | 71.8% | 1.4% | 49.6% | 4.8% | 68.0% | 1.4% |
| Sensitivity analysis:<br>Moderna vaccine initial<br>VE (95% CI) | 30.6%,<br>83.7% | Equal<br>to base<br>case | 48.8%,<br>84.5% | Equal<br>to base<br>case | 18.3%,<br>80.8% | Equal<br>to base<br>case | 41.8%,<br>82.4% | Equal<br>to base<br>case |
| Sensitivity analysis:<br>RR 95% CI (Moderna<br>vs. Pfizer-BioNTech<br>vaccines) for initial VE | Equal to base case |  |  |  | 42.8%,<br>55.8% | Equal<br>to base<br>case | 64.3%,<br>70.9% | Equal<br>to base<br>case |
| Sensitivity analysis:<br>Fall 2023 vaccine<br>waning | Equal<br>to<br>base<br>case | 3.1%,<br>6.8% | Equal<br>to<br>base<br>case | 0.6%,<br>2.4% | Equal<br>to<br>base<br>case | 3.1%,<br>6.8% | Equal<br>to<br>base<br>case | 0.6%,<br>2.4% |
| Sensitivity analysis:<br>differential waning for<br>Moderna & Pfizer-BioNTech vaccines* | Equal to base case |  |  |  | Equal<br>to<br>base<br>case | 5.6% | Equal<br>to<br>base<br>case | 1.6% |

\*The monthly waning rate for the Pfizer-BioNTech vaccine was adjusted so that the relative risk of infection and hospitalization between Moderna and Pfizer-BioNTech vaccines (0.85 and 0.88, respectively) was maintained for each month of the time horizon.
VE: vaccine effectiveness; CI: confidence interval; RR: relative risk;

### Infection consequences decision tree

The transition probabilities associated with the consequences of infection included in the infection consequence decision tree (Figure 3), are summarized in Table 4. Input data were obtained from targeted literature reviews and were estimated based on published sources, fee schedules, and databases. Where available, Canadian sources and data were used.

**Table 4.** Base case infection consequences decision tree probabilities.

| Age group | Hospitalization rates<br>(unvaccinated) given<br>infection, by age<br>(%)<sup>9,34,35*</sup> | Hospitalization locations of care, by age (%)<sup>29</sup> |  |  | In-hospital<br>mortality rates,<br>by age<sup>8,36†</sup> |
| --- | --- | --- | --- | --- | --- |
|  |  | No ICU or<br>ventilation | ICU | ICU with<br>ventilation |  |
| 18-19<br>years | 0.5% | 89.11% | 5.41% | 5.48% | 1.08% |
| 20-29<br>years | 1.1% | 91.81% | 4.07% | 4.12% | 1.68% |
| 30-39<br>years | 2.1% | 89.90% | 5.02% | 5.08% | 2.56% |
| 40-49<br>years | 2.8% | 86.32% | 6.80% | 6.88% | 5.58% |
| 50-59<br>years | 4.9% | 85.21% | 7.35% | 7.44% | 6.98% |
| 60-69<br>years | 10.8% | 86.51% | 6.70% | 6.79% | 9.54% |
| 70-79<br>years | 24.8% | 90.77% | 4.59% | 4.64% | 12.19% |
| 80+ years | 30.1% | 95.82% | 2.08% | 2.10% | 20.64% |

\* Adjusted for Omicron<sup>35</sup> and immunocompromised status<sup>9</sup>
† Only applied to hospitalized patients
ICU: Intensive care unit

Infected individuals either require hospitalization (inpatient care), or they do not. To inform the risk of hospitalization in those without protection from COVID-19 vaccination, the model requires the risk of hospitalization given infection in the unvaccinated population. As current Canadian data are not disaggregated by vaccination status, the age-specific risk of hospitalization given infection from the pre-vaccination period (July-December 2020) was first used and calculated from PHAC data using number of infections and hospitalizations by age.^34^ These data were then adjusted to the omicron period using the RR (0.58) of hospitalization between the omicron and delta period from a study by Wang et al., (2022).^35^ These age-specific risks of hospitalization during omicron given infection were then further adjusted to the immunocompromised population using the RR (1.94) of hospitalization between moderately and severely immunocompromised individuals and those not considered at high risk of severe outcomes during omicron, using data from a Canadian study by Bahremand et al (2023).^9^ Final model inputs are displayed in Table 4.

Hospitalization stay is categorized by the highest level of care required (general ward, ICU without ventilation, ICU with ventilation). The distribution, by age, were obtained from September 2022-August 2023 PHAC data.^29^. Deaths were only assumed to occur in those requiring hospitalization. Probabilities of death for the general population in those hospitalized, by age, were calculating by dividing the number of deaths by the number of hospitalizations from 2022-23 PHAC data.^36^ These values were increased in the IC population for all ages by applying the odds ratio for mortality between immunocompromised and immunocompetent individuals from Turtle et al. (2023).^8^ As data were not disaggregated by hospital location of care, probabilities were assumed to be the same for all.

All individuals with COVID-19 infection are subject to a “post-infection” cost and disutility toll representing longer-term cost impacts. ^37^ As the “post-infection” costs and disutility represented the average cost of all patients with a COVID-19 infection, it also included individuals who developed long COVID. Therefore, long COVID was not represented separately in the model.

### Resource use and costs

For the base case, resource use and costs associated with the treatment of infections and their consequences are considered. As coverage in both comparator arms are expected to be the same, and rates of adverse events are assumed to be similar and costs associated with adverse events are also excluded. Therefore, only resource use and costs incurred due to the treatment of COVID-19 infection (acute, post-infection, and from the societal perspective, productivity loss) are included. Acute infection resource use includes non-hospitalized (emergency department and physician visits) and hospitalizations due to acute infection.

Post-infection costs include ongoing care following the initial acute infection. Resource use was based on a Canadian retrospective database study on individuals’ post COVID-19 test. The incremental resource use in those with positive tests were calculated by comparing use by those with negatives tests, starting 56 days post test, and represented a year of resource use.^37^ Incremental resource use included hospitalization, outpatient medical encounters, home care, emergency department visits, and long-term care days. Costs for each resource were applied based on standard fee schedules.^37^

All costs were inflated to 2022 Canadian dollars using Statistics Canada’s Consumer Price Index.^38^. Cost inputs from the health care perspective are displayed in Table 5. Societal cost inputs are provided in the Supplementary Material.

**Table 5.** Cost inputs.

| Model parameter | Value | Assumptions |
| --- | --- | --- |
| Short-term infection: non-hospitalized | \$111 | Cost per outpatient visit<sup>39</sup> and emergency department visits,<sup>39,40</sup> adjusted for average number of outpatient visit (0.34) and emergency department visit (0.25) per infection.<sup>41</sup> |
| Hospitalization (per stay) |  |  |
| No ICU or ventilation | \$19,484 | Average cost April 2022-September 2022.<sup>42</sup><br>Cost for ICU with ventilation stay assumed same as ICU stay as data source did not differentiate the two. |
| ICU | \$54,672 | |
| ICU with ventilation | \$54,672 | |
| Post-Infection | \$513 | Reported resource use, including average number of outpatient visits, hospitalization days, and long-term care days.<sup>37-39,43</sup> Not disaggregated by hospital status. |
| **Quality-Adjusted Life-Years Lost** |  |  |
| Symptomatic infection, not hospitalized<sup>44,45</sup> | 0.003 | Applied to all symptomatic infections regardless of whether they were medically-attended or not. |
| Hospitalized, general ward<sup>46,47</sup> | 0.008 | The symptomatic infection, not hospitalized decrement is also added to the hospitalized |
| Hospitalized, ICU (no ventilator)<sup>46,47</sup> | 0.016 |  |
| Hospitalized, ICU with ventilator<sup>46,47</sup> | 0.030 | values to reflect symptoms prior to hospitalization |
| Post-infection, not hospitalized<sup>48</sup> | 0.0277 |  |
| Post-infection, hospitalized<sup>49</sup> | 0.122 |  |

ICU: Intensive care unit; QALY: quality-adjusted life-year

### Quality-adjusted life-years

Infections, hospitalizations, post-infections, and death are all associated with quality-adjusted life-year (QALY) decrements. The impact of the morbidity associated with COVID-19 infections is captured as the QALY decrements provided in Table 5.The estimated number of life-years lost due to premature COVID-19 mortality was calculated using the most recent Statistics Canada data on life expectancy by age.^50^ This was multiplied by the Canadian average population values for QALYs experienced by age.^51^ All future QALYs were discounted to present value at rate of 1.5%.^52^

### Scenario analyses and deterministic sensitivity analyses

In addition to the scenario and sensitivity analyses previously mentioned, analyses were conducted on incidence, hospitalization and mortality rates, costs, and QALY decrements. Where available, lower and upper 95% confidence intervals were used. All other values were varied by ± 25%. Value used for the included sensitivity analyses are presented in the Supplementary Material.

## RESULTS

In the base case analysis, assuming an IC population size of 894,580, there is expected to be 65,536 symptomatic infections, 7,151 hospitalizations and 1,133 deaths when vaccinating the Canadian IC adult population with the Moderna Fall 2023 vaccine from September 2023 – August 2024. This was achieved with 359,067 vaccinations administered between September 2023-January 2024. Given the expected lower VE of the Pfizer-BioNTech vaccine, the Moderna vaccine is expected to prevent 2,411 infections (3.5%), 275 hospitalizations (3.7%), and 47 deaths (4.0%) compared to vaccinating with Pfizer-BioNTech. Additionally, 330 more QALYs would be gained with the Moderna Fall 2023 vaccine, with over half being attributed to the prevention of death (Table 6). From the health care perspective, the prevented infections and hospitalizations with use of the Moderna Fall 2023 vaccine results in a $7.4 million saved in treatment costs; when productivity losses are included, $8.3 million in treatment cost savings are realized.

**Table 6.** Base case results.

|  | **Moderna Fall 2023**<br>**vaccine** | **Pfizer-BioNTech**<br>**Fall 2023 vaccine** | **Difference\*** |
| --- | --- | --- | --- |
| **Clinical Outcomes** |  |  |  |
| Number of vaccinations | 359,067 | 359,067 | 0 |
| **Cases** |  |  |  |
| Symptomatic infections | 65,585 | 67,996 | -2,411 |
| Hospitalizations | 7,151 | 7,426 | -275 |
| COVID-19 related deaths | 1,133 | 1,180 | -47 |
| **QALYs lost** |  |  |  |
| Morbidity | 2,587 | 2,682 | -96 |
| Mortality | 6,457 | 6,692 | -235 |
| **Total QALYS lost** | **9,044** | **9,374** | **-330** |
| **Economic Outcomes** |  |  |  |
| Short-term infection costs | \$164,397,246 | \$170,630,436 | -\$6,233,190 |
| Post-infection period | \$33,054,168 | \$34,266,464 | -\$1,212,296 |
| **Total Health care perspective** | **\$197,451,414** | **\$204,896,900** | **-\$7,445,486** |
| Productivity loss | \$39,488,494 | \$40,338,892 | -\$850,397 |
| **Total societal perspective** | **\$236,939,908** | **\$245,235,792** | **-\$8,295,884** |

\*Moderna Fall 2023 vaccine- Pfizer-BioNTech Fall 2023 vaccine
QALY: quality-adjusted life-year

Results from scenario analyses on incidence and vaccine uptake continued to find treatment cost savings and QALYs saved with the Moderna Fall 2023 vaccine compared to the Pfizer-BioNTech vaccine. When the incidence of COVID-19 infection was increased or decreased, the difference between the Moderna and Pfizer-BioNTech Fall vaccines for outcomes were increased or decreased by the same percentage. Increasing the Fall 2023 vaccine uptake in those aged 80 years and older to 80% increased costs saved and QALYs gained by the Moderna Fall vaccine compared to the Pfizer-BioNTech version by 9%, and increasing the uptake to 95% increased the costs saved and QALYs gained by 30%, as more IC individuals are vaccinated, and therefore contributing to the predicted benefits. Sensitivity analyses are presented in Figure 4. Tornado diagrams summarizing the sensitivity analyses for both costs saved and QALYs gained with the Moderna Fall vaccine compared to the Pfizer-BioNTech Fall vaccine are displayed. For ease of comparison, parameters for both diagrams are listed in decreasing order according to differences in cost saved. Parameters that did not have an impact on outcomes were excluded from the diagrams. In the base case, the starting VE against infection and hospitalization for Pfizer-BioNTech were calculated using the Wang et al. RRs,^20^ and the same waning rates were then applied to both vaccines. Costs and QALYs were most sensitivity to changes where the RR of infection and hospitalization was maintained between both vaccines for the entire analytic time horizon, by adjusting the waning rate of Moderna and Pfizer-BioNTech vaccines upward. This resulted in faster waning rates for the Pfizer-BioNTech vaccine. Next, model outcomes were sensitivity to the RR for infection and hospitalizations between the vaccines, with higher RRs resulting in lower costs and QALYs saved, and vice versa. The Moderna Fall vaccine initial VE was also a driver of costs and QALYs, with higher initial VEs resulting in smaller VE differences between the vaccines, and therefore a smaller difference in outcomes.

**Figure 4.**
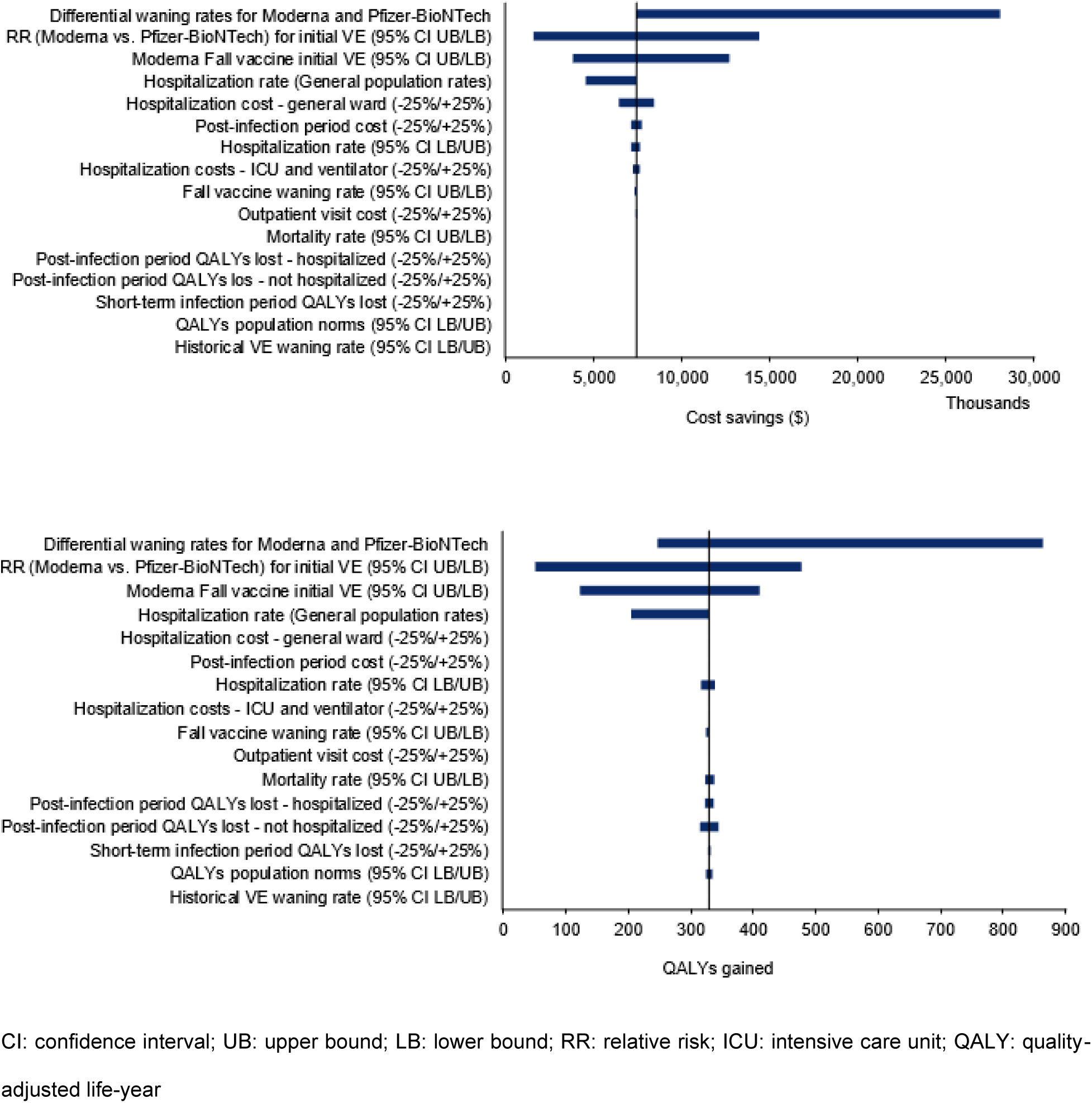
Sensitivity analyses tornado diagrams: cost and QALYs.

## DISCUSSION

This study estimated the difference in number of infections, hospitalization and death in the Canadian IC population over a one-year time horizon, with Moderna updated Fall 2023 vaccine administration compared to Pfizer-BioNTech updated Fall 2023 vaccine administration. Given the higher predicted VE of the Moderna Fall 2023 vaccine compared to the Pfizer-BioNTech vaccine, in a population size of 894,580 IC adults, vaccinating with the Moderna vaccine is predicted to prevent an additional 2,411 infections, 275 hospitalizations and 47 deaths, resulting in $7.4 million in health care cost savings, while gaining 330 QALYs. If productivity loss is included, the cost saving could be $8.3 million over the one-year time horizon.

The model is a simplification of the effects of vaccination on a population. It only accounts for direct protection of those receiving the vaccine, and only included the population of interest. It does not account for indirect protection through decreasing transmission in the general population. However, as the IC population is only a small sub-set of the entire Canadian population, vaccinating these individuals is unlikely to impact transmission in the general population.^53,54^

The SARS-CoV-2 virus is constantly evolving, and there is high uncertainty around future variants and incidence, and how responsive vaccines will be. Furthermore, as in many countries, many individuals no longer seek a medical diagnosis for their symptoms and there is under-reporting of COVID-19 infections in Canada. However, using the most recent information available on hospitalizations, we predicted the possible infection incidence for September 2022 to August 2023. We assumed that the incidence of infection for September 2023 to August 2024 will be the same as the previous year, however, it is impossible to know how COVID-19 will behave in this new post-pandemic era. The incidence of COVID-19 infection was calculated from the general population. While the IC population is at higher risk for more severe outcomes if they develop an infection, it is not known if the incidence of infection is different from the general population. As static models are linear by definition, a linear impact is seen on many parameters. For example, in the scenario analyses where incidence was increased/decreased by 25%, differences between the arms for costs saved and QALYs gained also increased/decreased by 25%.

Model results are highly sensitive to VE values of the Moderna and Pfizer-BioNTech Fall vaccines. While it is true that the VE values used in this analysis are assumptions and not based on real world data of the current XBB1.5 version of the vaccine, robust immunological response are induced by the vaccines and were in the same magnitude of increase compared to previous formulations against the intended variants.^55^ Therefore, the estimate of the Fall 2023 vaccines are based on real world data on previous versions of each vaccine. Despite the overall median follow-up duration of 2.46 months, VE estimates against hospitalization from Tseng et al., (2023) were used as it provided Moderna vaccine specific estimates during the time of BA.4/BA.5 dominance. This is in alignment with other modeling approaches.^56^ Previous studies have found higher VE values with Moderna vaccines compared to the Pfizer-BioNTech counterpart,^11–15,20^ including in other vulnerable populations, such as residents in nursing and retirement homes.^57^ The data used in this analysis to estimate Pfizer-BioNTech Fall 2023 vaccine VEs were obtained from a meta-analysis, which is considered the highest level of evidence in evidence-based medicine.^58^ Difference in the VEs between the vaccines may be due in part to different formulation, dosage, and delivery system. ^20^ In their Communicable Disease Control Manual, The British Columbia Centre for Disease Control lists the Moderna Fall 2023 vaccine as the preferred vaccine over the Pfizer-BioNTech version for IC individuals due to its potentially greater immune response,^9^ which is correlated with VE.^59,60^ If we expect similar VE outcomes as in previous version of the vaccine, then we can expect fewer infections, hospitalizations and deaths and lower COVID-19 treatment costs with the Moderna updated Fall 2023 vaccine.

Although there are study limitations due the lack of IC specific data, steps have been taken to adjust existing general population data, and determine the impact of uncertainty. Because data were unavailable on the VE against infection of prior Moderna formulations in the IC population, or on the relative VE for the general population compared to the IC population, we were unable to estimate the adjusted VE against infection for the Moderna updated Fall 2023 vaccine from the same source used to estimate the VE against hospitalization, and assumed the same VE against infection as mRNA-1273 against BA.1/BA.2 in the general population.^30^ Although different populations and time periods were used, the VE estimate against infection is conservative. If the VE of the Moderna updated Fall 2023 vaccine against infection was assumed to be lower in the IC population, then the estimated number of infections prevented compared to the Pfizer-BioNTech vaccine would be even larger (see Supplementary Material for results).

The population size was estimated using proportion estimated from the US, as suitable data were not available from Canada. Over or underestimating the population size will result in the total costs saved and QALYs gained to be over or underestimated, respectively.

Additionally, monthly VE waning rates were obtained from the general population. A study by Ferdinand et al. (2022), reported higher hospitalization waning rates over a six-month period in IC individuals during Delta/Omicron predominance.^61^ However, increasing the waning rates in sensitivity analysis had minimal effects on cost and QALYs as the waning rate was the same for both the Moderna and Pfizer-BioNTech Fall 2023 vaccines.

Although hospitalization and mortality rates were used for the Canadian general population, efforts were made to adjust these to the IC population by applying the increased risk observed in the IC population compared to immunocompetent individuals. In all sensitivity analyses surrounding these parameters, the Moderna Fall 2023 vaccine continued to result in cost savings and QALYs gained compared to the Pfizer-BioNTech Fall vaccine. Additionally, QALYs related to COVID-19 infection and hospitalizations were based on the general population, and were not adjusted due to lack of data. However, varying these values in sensitivity analyses had minimal impact on total QALYs gained.

The proportion of hospitalized IC individuals that required ICU admission was based on the general population data as Canadian data on the IC population was not available. However, a UK study by Evans et al., (2023) found the 2.8-4.7% of hospitalized IC individuals in 2022 required ICU admission compared to 2.1% overall. Based on PHAC data, the age-dependent rate of ICU admission given hospitalizations in the general population between July 2022-August 2023 ranged from 4.2%-10.9%. Although the UK and Canadian populations included are not comparable, the UK study suggests that there may be a higher proportion of ICU admissions in hospitalized patients compared to the general population. As such, given that the Moderna Fall 2023 vaccine is predicted to result in fewer hospitalization compared to the Pfizer-BioNTech Fall 2023 vaccine, the cost savings and QALYs estimated by the current study is likely to be underestimated.

Likewise, resource use and costs included in the analyses were obtained from the general population, and not IC individuals, resulting in conservative estimates. Preliminary findings comparing the two populations found that in the US, IC individual COVID-19 inpatient expenditures were 1.4 times greater than non-IC individuals.^62^ If healthcare expenditures up to 60 days post discharge were included, this value increases to 1.8 times greater. While the US and Canadian health care systems and costing are not comparable, this study provides evidence that IC individuals may require a higher volume of health care, and that the cost inputs used in the current analysis may therefore be underestimated. Increasing these cost inputs would increase the cost savings by the Moderna Fall 2023 vaccine compared to the Pfizer-BioNTech vaccine, as confirmed in the cost sensitivity analyses. Therefore, the limitations in resource use and cost values used in the base case result in extremely conservative estimates of the costs saved by use of the Moderna Fall 2023 vaccine.

Various other sensitivity analyses were conducted on the VE estimates, including the difference in VE between the vaccines, and varying the monthly waning rate. Still, in every analysis, overall results remained the same, with the Moderna Fall 2023 vaccine saving costs and gaining QALYs compared to the Pfizer-BioNTech Fall 2023 vaccine. Cost savings varied from $1.6 million, where the relative risk of infection or hospitalizations between the two vaccines was closer to 1, resulting in a smaller difference in VE values, to $28.1 million, when the waning rate of Pfizer-BioNTech was set higher. QALYs gained ranged from 71 to 1,227 (resulting from same sensitivity analyses mentioned for cost savings). These findings were largely driven by the changes in the number of hospitalizations.

The model only allowed those who completed primary series vaccination to receive a Fall 2023 vaccine, as was the case for the bivalent booster. The current NACI guidelines recommend the XBB.1.5-containing formulations for those previously vaccinated. As the vaccines are also authorized by Health Canada for those not previously vaccinated, NACI states that they may be used to initiate primary series. However, recommendations for this population will be forthcoming in the next months.^2^ Additionally, as the historical ICES database coverage data from ICES found that only 15% of IC individuals aged 60 and above did not complete primary series,^24^ it is unlikely that vaccination of a small proportion of individuals with primary series would change model results. Saskatchewan has indicated that moderately to severely IC individuals who have not received primary series be vaccinated with multiple doses of the Fall 2023 vaccines, with immunocompetent individuals requiring one.^63^

In conclusion, this study conservatively estimated the potential benefits of vaccinating the Canadian IC adult population with the Moderna updated Fall 2023 vaccine instead of the Pfizer-BioNTech Fall 2023 vaccine. If the Moderna and Pfizer-BioNTech Fall 2023 vaccines are well-matched to the dominant variants and protect against infection and hospitalizations similar to previous versions of the vaccines, using the Moderna Fall 20203 vaccine could result in substantial health care and societal cost savings, as well as QALYs gained.

## Declarations

### Ethics approval and consent to participate

Not applicable

### Consent for publication

Not applicable

### Availability of data and materials

The datasets used and/or analyzed during the current study are available from the corresponding author on reasonable request.

### Authors’ contributions

Conceptualization, AL, MK, MM, KaJ, NV; Methodology, AL, KaJ, MK, MM, EB, and NV; Software, MM; Validation, AL and MK; Formal Analysis, AL and KF; Writing – Original Draft Preparation, AL and MK; Writing – Review & Editing, KaJ, KeJ, EB, NV; Funding Acquisition, KaJ, EB, NV

## Acknowledgements

Not applicable

## Author’s information (optional)

Not applicable

