## Supplementary material for "Clinical and Economic impact of updated Fall 2023 COVID-19 vaccines in the Immunocompromised Population in Canada"

### CONTENTS

Population size 3

Incidence 4

Productivity loss costs 6

List of sensitivity analyses 9

Base case results 11

Sensitivity analyses results 12

### LIST OF TABLES

Table 1. Canada immunocompromised population size, by age 3

Table 2. Hospitalization rate in general population, given infection 4

Table 4. Productivity loss inputs 7

Table 5. List of deterministic sensitivity analyses 9

Table 6. Base case results 11

### LIST OF FIGURES

Figure 1. COVID-19 projected hospitalization, per 100,000 population, by age 5

Figure 2. COVID-19 projected incidence, by age group 6

Figure 3. Sensitivity analyses tornado diagrams, with parameters ordered by differences in cost saved 12

#### Population size

A Statistics Canada study report that 14% of individuals age 15+ responded “Yes” to whether they have a compromised immune system, however, further information on severity was not collected.[1] This is in contrast to an estimate from a US study, that asked additional questions to gauge the accuracy and severity of the response.[2] To estimate the population size in Canada, it was assumed that the proportion of IC adults is the same as in the USA (2.7%). To estimate the number of IC adults in Canada by age, values estimated by age group from the Statistics Canada report were adjusted down based on overall levels found in the USA report, and multiplied by the 2022 population in Canada, by age. This is displayed in Table 1.

**Table 1. Canada immunocompromised population size, by age**

| Age group | Immunocompromised population size |
|---|---|
| 18-19 years | 9,836 |
| 20-29 years | 60,448 |
| 30-39 years | 95,125 |
| 40-49 years | 115,233 |
| 50-59 years | 165,855 |
| 60-69 years | 188,935 |
| 70-79 years | 160,671 |
| 80+ years | 98,477 |

#### Incidence

The current incidence of COVID-19 in Canada is unknown due to the lack of mandatory testing and reporting. However, hospitalization data are still available. Therefore, to estimate the incidence, a modified version of the static model was used to estimate incidence given hospitalization rates, through a calibration process.

The rate of hospitalization given infection was calculated in the unvaccinated general population, based on data from July – December 2020, a period where both number of infections and hospitalizations in Canada were reported, and when vaccinations were not yet available.[3] These rates were then adjusted down to account for the less sever Omicron variant[4]. Resultant hospitalization rates for the general population are displayed in Table 2.

**Table 2. Hospitalization rate in general population, given infection.**

| Age group | Hospitalization rates |
|---|---|
| 18-19 years | 0.28% |
| 20-29 years | 0.28% |
| 30-39 years | 0.56% |
| 40-49 years | 1.07% |
| 50-59 years | 1.42% |
| 60-69 years | 2.54% |
| 70-79 years | 5.58% |
| 80+ years | 12.80% |

Hospitalization rates in the general population from September 2023-August 2024 were assumed to be the same as those observed between September 2022-August 2023. At time of analysis, data up to June 2023 were available, with July-August 2023 data being incomplete.[5] July and August 2023 data were therefore estimated by applying the ratio of June 2023 to June 2022 rates, and applying them to the July-August 2022 data. These target hospitalization rates, which the incidence rates were calibrated against, are displayed in Figure 1.

**Figure 1. COVID-19 projected hospitalization, per 100,000 population, by age**

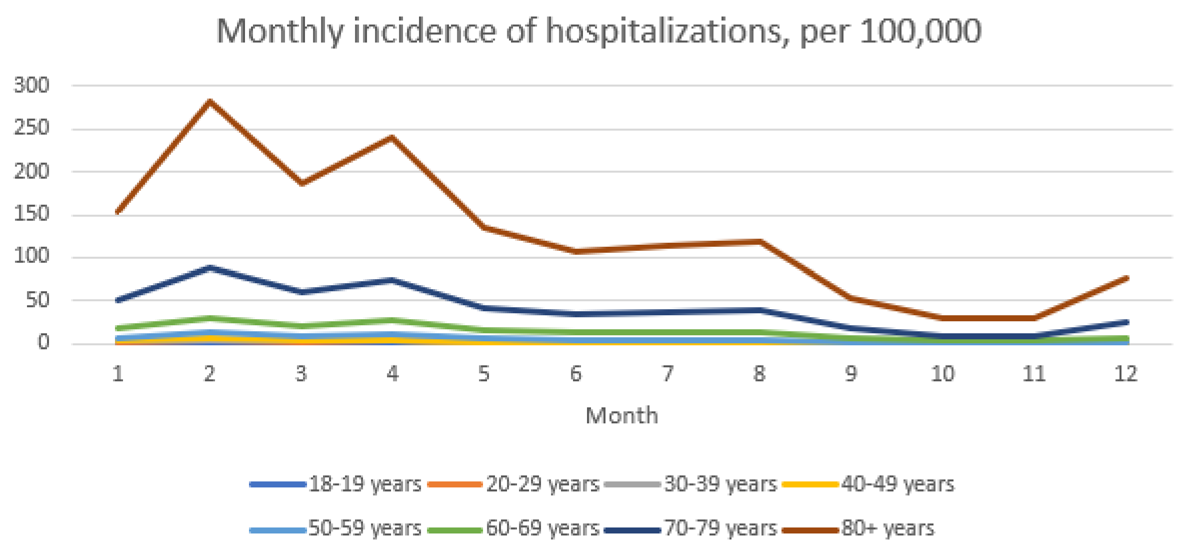


The projected COVID-19 infection incidence for September 2023-August 2024, by age group, is provided in Figure 2.

**Figure 2. COVID-19 projected incidence, by age group**

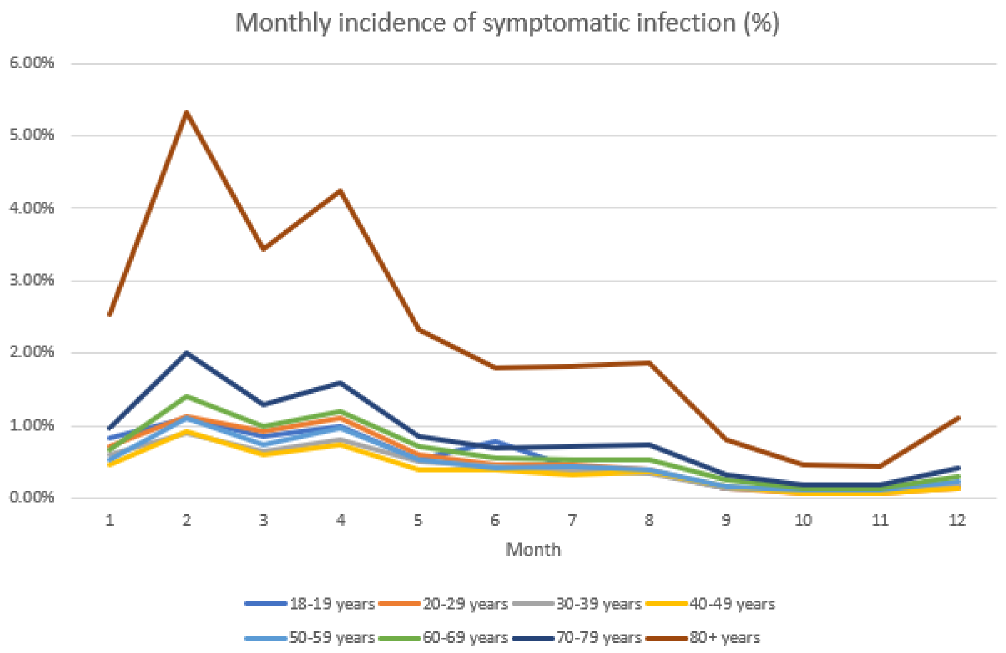


#### Productivity loss costs

The societal perspective includes all costs from the health care perspective, as well as costs due to productivity loss. The model includes productivity loss costs due to acute infection and hospitalizations. As an equal number of vaccinations was assumed regardless of vaccine administered, time loss due to administration was not included.

Productivity loss due to non-hospitalized infection was assumed to be 5 days. To adjust for non-work days, the value was decreased by 2/7 to account for the weekend. Productivity loss due to hospitalizations were based on the length of stay in hospital[6] and were also adjusted for non-working weekends. They were multiplied by the average daily wage in Canada,[7] assuming an 8-hour work day, and adjusting for labour force participation, by age.[8] Productivity loss inputs are displayed in Table 3.

#### Table 3. Productivity loss inputs

| Variable | Mean value | Source |
|---|---|---|
| Labour force participation rate | | |
| 18-19 years | 51.60% | Statistics Canada.[8] |
| 20-29 years | 82.63% | |
| 30-39 years | 89.00% | |
| 40-49 years | 89.41% | |
| 50-59 years | 81.69% | |
| 60-69 years | 43.65% | |
| 70-79 years | 8.00% | |
| 80+ years | 8.00% | |
| Daily wage | $255.68 | Statistics Canada.[7] Assume 8-hour work day |
| Time loss (days) | | |
| Not hospitalized | 3.6 | Assumption. Isolate until symptoms improving for 24 hrs (48 hrs if nausea, vomiting and/or diarrhea). Assumed 5 days adjusted for non-working weekends. |
| Hospitalized – general ward | 11.5 | Canadian Institute for Health Information[6] |
| Hospitalized ICU | 18.9 | Canadian Institute for Health Information[6] |
| Hospitalized ICU with ventilator | 18.9 | assume same as ICU |
| Total time loss for hospitalization* | | *Estimates include time loss for symptomatic infection, not hospitalized to reflect time with symptoms prior to |

| | | |
|---|---|---|
| | | hospitalization, and time loss associated with the hospitalization length of stay |
| No ICU or ventilator | 15.1 | |
| ICU only | 22.4 | |
| Ventilator | 22.4 | |

ICU, intensive care unit

#### List of sensitivity analyses

Deterministic sensitivity analyses and values varied for each analysis are displayed in Table 4. Where available, 95% confidence intervals were used. All other values were varied by ±25%.

##### Table 4. List of deterministic sensitivity analyses

| Parameter | Base-Case | Range |
|---|---|---|
| Moderna Fall vaccine initial VE | Tseng et al., for hospitalization,[9]<br>Pratama et al., for infection[10] | 95% CI |
| RR between Moderna and Pfizer-BioNTech Fall Vaccine | Wang et al.,[11] | 95% CI |
| Waning (Fall 2023 vaccine) | Higdon et al.,[12] | 95% CI |
| Differential waning between Moderna and Pfizer Fall vaccine | Waning rates for both set to Higdon et al.,[12] | Waning rates for Pfizer-BioNTech adjusted so that the RR observed between Moderna and Pfizer-BioNTech from Wang et al.[11]) are maintained |
| Historical VE | | 95% CI |
| Hospitalization rate | RR for IC population applied to general population to inflate values | General population hospitalization rates |
| | | 95% CI |
| Mortality rate | | 95% CI |
| Cost per hospitalization - general ward | $19,484 | ±25% |
| Cost per hospitalization - ICU and ventilator | $54,672 | ±25% |

| | | |
|---|---|---|
| Cost per outpatient visit | $111 | ±25% |
| Population norm utilities | From Guertin et al.[13] | 95% CI |
| Short term infection QALY decrement | 0.0026 | ±25% |
| Post-infection cost | From McNaughton et al.[14] | ±25% |
| Post-infection QALYs lost - not hospitalized | 0.028 | ±25% |
| Post-infection QALYs lost - hospitalized | 0.122 | ±25% |

CI, confidence interval, VE, vaccine effectiveness; RR, relative risk; ICU, intensive care unit; QALY, quality-adjusted life-year

#### Base case results

Disaggregated base case results are displayed in Table 5.

##### Table 5. Base case results

| | **Moderna Fall 2023 vaccine** | **Pfizer-BioNTech Fall 2023 vaccine** | **Difference*** |
|---|---|---|---|
| **Clinical Outcomes** | | | |
| Number of vaccinations | 359,067 | 359,067 | 0 |
| **Cases** | | | |
| Symptomatic infections | 65,585 | 67,996 | -2,411 |
| Hospitalizations | 7,151 | 7,426 | -275 |
| COVID-19 related deaths | 1,133 | 1,180 | -47 |
| **QALYs lost** | | | |
| Morbidity | 2,587 | 2,682 | -96 |
| Mortality | 6,457 | 6,692 | -235 |
| **Total QALYS lost** | 9,044 | 9,374 | -330 |
| **Economic Outcomes** | | | |
| **Short-term infection costs** | | | |
| Not hospitalized | $6,479,025 | $6,715,819 | -$236,794 |
| Hospitalized | $157,918,221 | $163,914,617 | -$5,996,396 |
| **Post-infection period** | | | |
| Not hospitalized | $29,967,993 | $31,063,257 | -$1,095,264 |
| Hospitalized | $3,086,175 | $3,203,207 | -$117,032 |
| **Total Health care perspective** | $197,451,414 | $204,896,900 | -$7,445,486 |
| Productivity loss | $39,488,494 | $40,338,892 | -$850,397 |
| **Total societal perspective** | $236,939,908 | $245,235,792 | -$8,295,884 |

*Moderna Fall 2023 vaccine- Pfizer-BioNTech Fall 2023 vaccine

QALY: quality-adjusted life-year

#### Sensitivity analyses results

Full results for economic (health care costs) and clinical (infections, hospitalizations, deaths, QALYs) outcomes from the sensitivity analyses are displayed in

Figure 3. As in the main manuscript, for comparison purposes, all tornado diagram parameters are ordered in decreasing order according to differences in cost saved.

**Figure 3. Sensitivity analyses tornado diagrams, with parameters ordered by differences in cost saved**

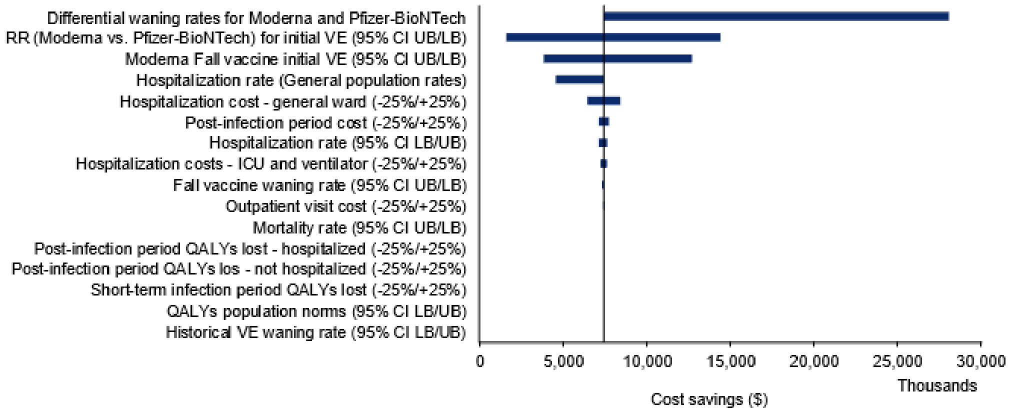


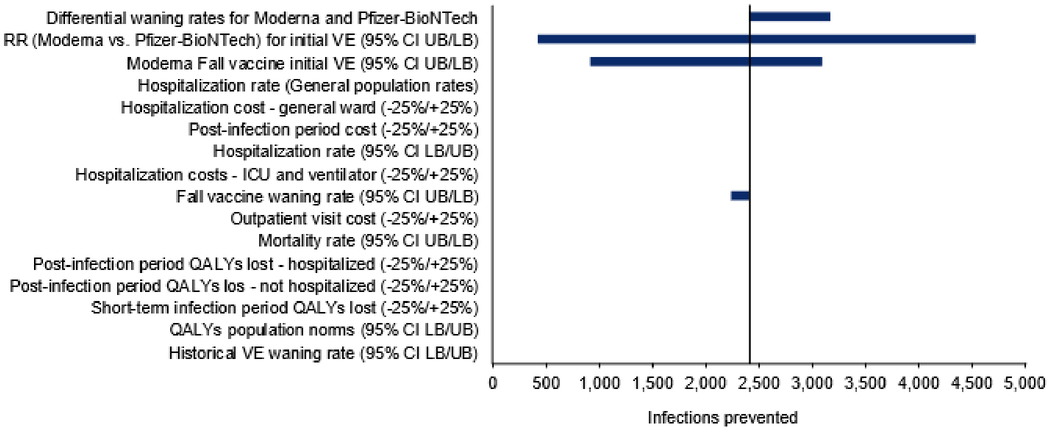

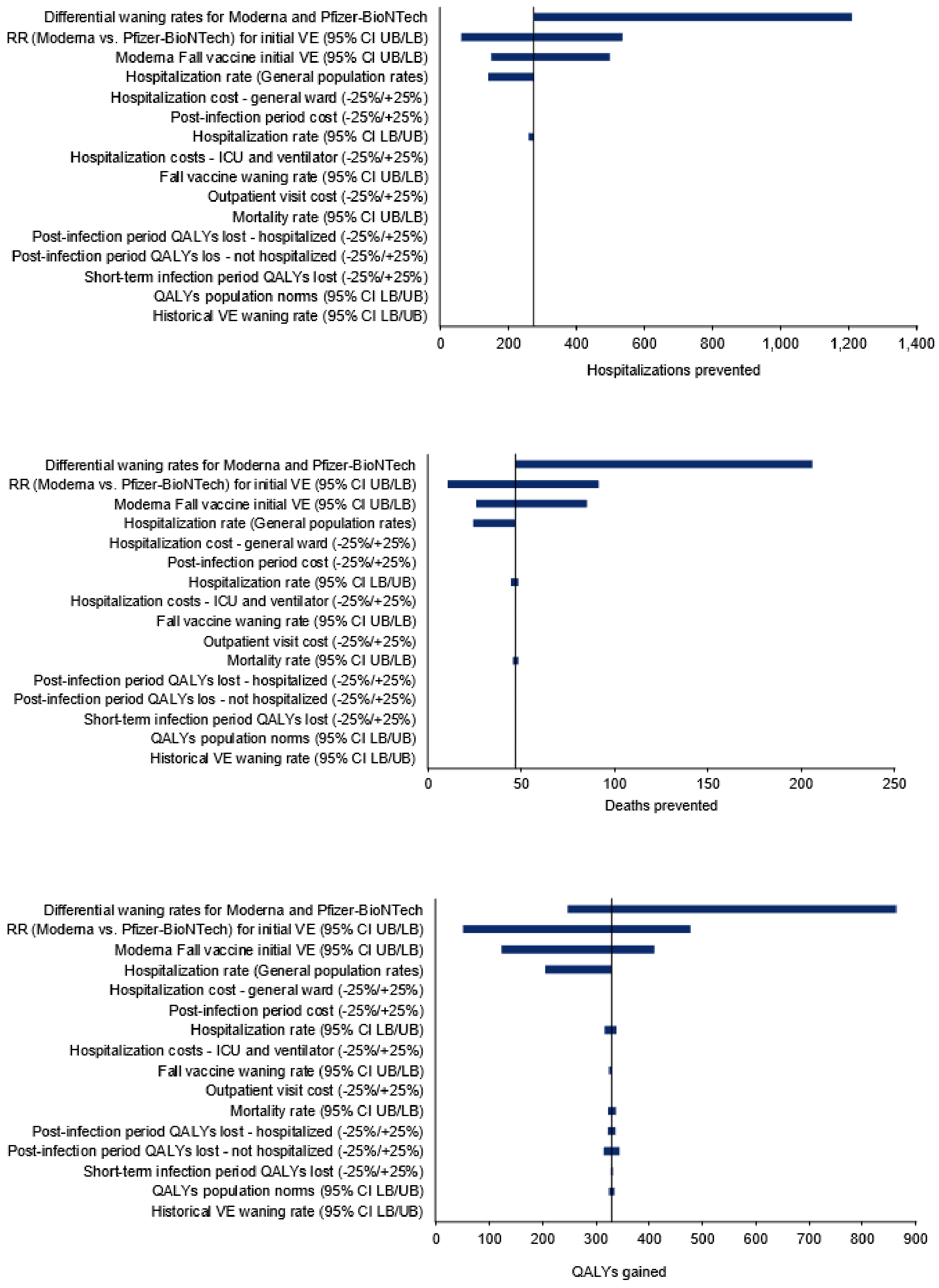


CI: confidence interval; UB: upper bound; LB: lower bound; RR: relative risk; ICU: intensive care unit; QALY: quality-adjusted life-year
